# Reproducibility of Genetic Risk Factors Identified for Long COVID using Combinatorial Analysis Across US and UK Patient Cohorts with Diverse Ancestries

**DOI:** 10.1101/2025.02.04.25320937

**Authors:** J Sardell, M Pearson, K Chocian, S Das, K Taylor, M Strivens, R Gupta, A Rochlin, S Gardner

**Author notes:** Joint last authors (A Rochlin, S Gardner).

## Abstract

**Background:** Long COVID is a major public health burden causing a diverse array of debilitating symptoms in tens of millions of patients globally. In spite of this overwhelming disease prevalence and staggering cost, its severe impact on patients’ lives and intense global research efforts, study of the disease has proved challenging due to its complexity. Genome-wide association studies (GWAS) have identified only four loci potentially associated with the disease, although these results did not statistically replicate between studies. A previous combinatorial analysis study identified a total of 73 genes that were highly associated with two long COVID cohorts in the predominantly (>91%) white European ancestry Sano GOLD population, and we sought to reproduce these findings in the independent and ancestrally more diverse All of Us (AoU) population.

**Methods:** We assessed the reproducibility of the 5,343 long COVID disease signatures from the original study in the AoU population. Because the very small population sizes provide very limited power to replicate findings, we initially tested whether we observed a statistically significant enrichment of the Sano GOLD disease signatures that are also positively correlated with long COVID in the AoU cohort after controlling for population substructure.

**Results:** For the Sano GOLD disease signatures that have a case frequency greater than 5% in AoU, we consistently observed a significant enrichment (77% - 83%, *p* < 0.01) of signatures that are also positively associated with long COVID in the AoU cohort. These encompassed 92% of the genes identified in the original study. At least five of the disease signatures found in Sano GOLD were also shown to be individually significantly associated with increased long COVID prevalence in the AoU population. Rates of signature reproducibility are strongest among self-identified white patients, but we also observe significant enrichment of reproducing disease associations in self-identified black/African-American and Hispanic/Latino cohorts. Signatures associated with 11 out of the 13 drug repurposing candidates identified in the original Sano GOLD study were reproduced in this study.

**Conclusion:** These results demonstrate the reproducibility of long COVID disease signal found by combinatorial analysis, broadly validating the results of the original analysis. They provide compelling evidence for a much broader array of genetic associations with long COVID than previously identified through traditional GWAS studies. This strongly supports the hypothesis that genetic factors play a critical role in determining an individual’s susceptibility to long COVID following recovery from acute SARS-CoV-2 infection. It also lends weight to the drug repurposing candidates identified in the original analysis. Together these results may help to stimulate much needed new precision medicine approaches to more effectively diagnose and treat the disease.

This is also the first reproduction of long COVID genetic associations across multiple populations with substantially different ancestry distributions. Given the high reproducibility rate across diverse populations, these findings may have broader clinical application and promote better health equity. We hope that this will provide confidence to explore some of these mechanisms and drug targets and help advance research into novel ways to diagnose the disease and accelerate the discovery and selection of better therapeutic options, both in the form of newly discovered drugs and/or the immediate prioritization of coordinated investigations into the efficacy of repurposed drug candidates.

## Introduction

Post COVID condition, commonly known as long COVID or PASC (post-acute sequelae of SARS-CoV-2 infection), is a debilitating chronic condition that develops following a SARS-CoV-2 infection in around 5-15% of patients^1^. The global prevalence of long COVID is estimated at least 65 million people^2^ and is increasing annually. It’s estimated to now have a cumulative global incidence of over 400 million individuals and cost over $1 trillion (or 1% of global GDP) annually^1^, which causes a long-lasting and profound impact on patients’ lives and healthcare systems and has created a major public health issue^3^.

‘Long COVID’ is a term originally defined by patients to describe the post-acute and long-term health effects of COVID-19^4,5^ and the highly variable symptoms associated with the condition. The frequency and severity of SARS-CoV-2 infections appears to be correlated with increased risk of developing long COVID^6^.

Long COVID patients have reported a diverse array of symptoms across multiple organ systems^7^ with the most common being post-exertional malaise^8^, dysautonomia^9^, cognitive dysfunction^10^, mood disturbances^11^ and respiratory problems^12^. Many of these symptoms and signs are also observed in other complex neuroimmune disorders such as myalgic encephalomyelitis/chronic fatigue syndrome (ME/CFS)^13,14,15^, postural orthostatic tachycardia syndrome (POTS)^16,17^ and fibromyalgia^18,19^, all of which, like long COVID, disproportionately affect women^20^. To advance our understanding of the pathophysiological mechanisms underlying these shared clinical manifestations, it is important to have a deeper understanding of the genetic similarities between long COVID and other neuroimmune conditions. This effort is hampered, as most of these diseases, like long COVID, are highly complex and have been intractable for existing genomic analysis approaches.

More than four years following the global COVID-19 outbreak, patients still often struggle to obtain a long COVID diagnosis as agreement on the definition of the disease remains elusive beyond self-reported persistence of a wide range of symptoms. Governments also find evaluating its prevalence and setting public health policy difficult due to absence of a clear and consistent definition of the disease^21,22^. There are currently no recognized laboratory diagnostic tests or disease modifying therapies for long COVID. Research into the biological mechanisms of the disease is hindered by the variability in study designs, lack of reproducible findings across patient populations, and challenges in accurately capturing the heterogenous clinical phenotypes of patient cohorts^23^. A definitive biological explanation of some of the factors causing and defining the disease and a test encompassing these is urgently required to overcome this.

Only a few preliminary GWAS for long COVID have been published to date ^24,25,26^, likely due to the challenges of assembling a sufficiently powered patient cohort and the studies’ consequently limited findings. A study by the COVID-19 Host Genetics Initiative (HGI) identified only a single significant locus (*FOXP4*) from an analysis of 6,450 long COVID cases and over 1 million population controls aggregated from multiple cohorts^25^. Another recent meta-analysis of over 53,000 cases and 120,000 controls from 23andMe identified three significant loci (*HLA-DQA1–HLA-DQB*, *ABO* and *BPTF–KPAN2–C17orf58*)^26^. The effect sizes of the latter three loci reproduced in the HGI cohort but the associations were not significant, likely due to limited statistical power even in such a large cohort. The reported association between *FOXP4* and long COVID did not reproduce in the 23andMe data.

### Combinatorial Analytics for Complex Diseases

Combinatorial analytics has been more successful than GWAS in identifying key genetic risk factors that capture the complex biology of similarly multifactorial and heterogenous diseases like ME/CFS, generating more mechanistic insights and reproducible findings across cohorts^27^.

The combinations of genetic variants (‘disease signatures’) identified by combinatorial analyses capture both the linear and non-linear effects of interactions between multiple genes. They can be used to identify individual patients who have specific disease signatures, enabling the identification of associations between the disease signatures associated with a mechanism and the symptoms presented by patients with those disease signatures. These can improve our understanding of complex diseases beyond the single SNP associations identified by GWAS^28,29^ and creates opportunities for clinically actionable diagnostic tests and the targeted trials of multiple drug repurposing candidates to provide clinical benefit to specific patient cohorts.

### Aims of Study

The PrecisionLife combinatorial analytics platform was previously used to identify disease signatures for Severe and Fatigue Dominant long COVID cohorts derived from the Sano Genetics’ long COVID GOLD (Sano GOLD) study, and to highlight the biological similarities and differences between these two patient populations^30^. At the same time, combinatorial analysis was also undertaken on a General long COVID cohort encompassing all patients with a broader (and potential less-reliable) definition of the disease. The General cohort’s results were not described in the original publication, which instead focused on the most well phenotyped cases.

The Severe cohort in this study was comprised of cases who self-reported the greatest variety and severity of symptoms, while the Fatigue Dominant cohort was comprised of cases who self-reported predominantly fatigue-associated long COVID symptoms. The study identified a total of 73 genes that were highly associated with at least one of these long COVID populations. Of these genes at the time of publication, 9 genes were linked to acute COVID-19, 14 genes were differentially expressed in a previous transcriptomic analysis of long COVID patients^31^ and 9 genes were found that had been associated with ME/CFS in the previous combinatorial analysis of this disease^27^.

In this study, we assessed the findings of all three of the original long COVID combinatorial analyses of the Sano GOLD cohorts in an independent, more ancestrally diverse patient population. We used genomic, clinical, and questionnaire data from the All of Us (AoU) population^32^ to generate a long COVID cohort (using ICD-10 code U09.9) and evaluated the reproducibility of the findings from the original Sano GOLD study. We investigated the genes and mechanisms underlying the reproducible disease signatures, and evaluated the clinical phenotypes associated with each.

## Materials and Methods

### Generation of long COVID cohorts

For this study, we identified a cohort of long COVID patients and matching controls from the AoU dataset (accessed on December 10^th^ 2024). AoU provides data^33^ for nearly 850,000 American participants, including genomic data derived from the Illumina Global Diversity Array (GDA)^34^ (n=312,925), electronic health records (EHR, n=254,700), health questionnaires (n=412,220), and COPE COVID-19 survey (n=100,220)^35^. The AoU dataset was designed to capture data for a diverse group of individuals, including non-European ancestry groups often underrepresented in genomic datasets, and the cohort selected for this study reflects this diversity.

The baseline long COVID cohort was created by selecting all 458 individuals with GDA genotyping data who have a diagnosis of long COVID, using ICD-10 code U09.9 (post-acute COVID-19). We note that this criterion, which implies a prevalence of long COVID less than 0.2%, almost certainly excludes many patients with long COVID based on published estimates of long COVID prevalence of between 6.9% to 14%^36,37,38^.

The control cohort was generated by selecting individuals with GDA genotyping data who have evidence of SARS-CoV-2 infection, either based on a reported positive COVID-19 test in the COPE COVID-19 survey (n=3,615) or presence of ICD-10 codes B97.21 or U07.1 (n=17,024). We excluded individuals with long COVID based on ICD-10 code U09.9 as well as any individual with a history of symptomatic phenotypes consistent with long COVID or other post-viral fatigue syndromes (see Supplemental Table 1). Applying these criteria, our maximum control population included 9,774 individuals.

We used the sex-imputation functionality of PLINK^39^ to identify the genetic sex of each of the individuals in the full GDA dataset. 2.9% of total samples could not be reliably identified as male or female and were excluded from the study. 57.6% were identified as female and 39.5% were identified as male, which broadly agrees with the self-reported distribution of sex at birth from the AoU demographics questionnaire (59% female, 39% male, 2% skip/unknown).

### Case and control matching using stratified sampling

To create a balanced dataset and reduce potential confounding effects of population substructure, we created a subset of controls that match the demographic distribution (i.e., sex and self-reported race/ethnicity) of the long COVID cases. We used genetic sex inferred by PLINK (using the command `--check-sex`) as well the answers to the demographics survey on self-reported race and ethnicity to split the cohort into subgroups, and we compared the percentage of the baseline cases and potential controls that fall in each category. The results showed that some subgroups were over-or under-represented in cases vs controls, e.g. white, female, non-Hispanics accounted for 38.5% of cases but only 29.7% of controls.

We adjusted our long COVID cohort by removing all individuals whose sex was undetermined during PLINK sex-imputation. We also removed all individuals whose self-reported race and/or ethnicity was coded as “None of these/I prefer not to answer/PMI:Skip” as these demographics do not allow accurate ‘matching’ with the control population. This created a final long COVID case population of 413 individuals (see Table 1 for demographic distribution).

**Table 1.** The distribution by genetic sex, self-reported race, and self-reported ethnicity of cases and controls in the final All of Us long COVID cohort.

| Demographic | Subgroup | Final cases %<br>(n=413) | Baseline Controls %<br>(n=8,683) | Stratified Sampled Controls % (n=4,130) |
| --- | --- | --- | --- | --- |
| Genetic sex | F | 69.7% | 65.2% | 68.3% |
| Genetic sex | M | 30.3% | 34.8% | 31.7% |
| Self-reported race | Asian or None Indicated | 15.7% | 24.9% | 17.0% |
| Self-reported race | Black or African-American | 18.6% | 19.5% | 19.5% |
| Self-reported race | White | 65.6% | 55.6% | 63.5% |
| Self-reported ethnicity | Hispanic or Latino | 17.2% | 26.4% | 18.5% |
| Self-reported ethnicity | Not Hispanic or Latino | 82.8% | 73.6% | 81.5% |

The set of potential controls allowed us to create a final study cohort with a 1:10 case:control ratio and similar demographic splits in the case and control sub-cohorts. We used a probabilistic function to apply a stratified sampling technique using granular subgroups based on three demographic values (as illustrated in Table 2) to the baseline potential controls and match the distribution of the demographic subgroups in the long COVID cases as closely as possible.

**Table 2.** Two examples of how stratified sampling balances the frequency of granular subgroups (full breakdown is not available due to reporting restrictions imposed by AoU for rare subgroups).

| Sex | Self-reported race | Self-reported ethnicity | Final Cases (%) | Baseline Controls (%) | Stratified Sampled Controls (%) |
| --- | --- | --- | --- | --- | --- |
| F | Black or African-American | Not Hispanic or Latino | 14.5 | 13.0 | 14.7 |
| F | White | Not Hispanic or Latino | 42.6 | 33.2 | 40.2 |

Prior to sampling, we also removed any age-based outliers from the control cohort (i.e., any individuals whose age was outside the range of ages in the long COVID case cohort). Additional information on the demographic breakdown of cases and controls, including prevalence of comorbidities is included in Supplemental Table 2. The concordance between the self-reported demographic data and the AoU genetic ancestry predictions^33^ was very high (88.0-99.4% for matched groups) in the study cohort (Supplemental Table 3).

We used principal component analysis (PCA) to model any remaining population substructure within the AoU study cohort. We first removed all SNPs that are associated with the sex chromosomes or the MHC region on chromosome 6 or that have minor allele frequency less than 0.05. We then conducted LD-pruning in PLINK 1.9 (`--indep-pairwise 50 5 0.2`) before generating genetic PCs using the PLINK `--pca` command, as recommended elsewhere^40,41,42^.

We selected the top 5 PCs for use in our analyses based on the associated eigenvalues (Supplemental Table 3)

### Long COVID Disease Signatures

We previously identified long COVID associated disease signatures in two patient cohorts derived from the Sano GOLD study cohort, as described in the original Taylor *et al*. (2023) paper^30^, and a third unreported patient cohort using a broader definition of the disease (Supplemental Table 4). This resulted in:

1. 1,188 signatures mapped to 43 genes, from a ‘Severe’ cohort of patients who reported the greatest variety and severity of long COVID symptoms.
2. 1,435 signatures mapped to 35 genes, from a ‘Fatigue Dominant’ cohort of patients who reported predominantly fatigue-associated long COVID symptoms.
3. 6,445 signatures mapped to 165 genes, from a ‘General’ cohort of patients who reported they were still suffering continuation or development of new symptoms 12 weeks after the initial SARS-CoV-2 infection, with these symptoms lasting for at least 2 months with no other explanation.

In contrast to the diverse AoU American study cohort, the Sano GOLD study cohort was comprised of British patients of predominantly white European ancestry (>91% of the cohort), with Asian ancestry (∼4%) as the largest non-white European demographic.

### Evaluating Enrichment of Reproducing Long COVID Disease Signature in AoU Cohort

We used a logistic regression approach to evaluate the disease association of each of the previously identified disease signatures in the AoU study population. Individuals were coded as 1 if they possessed the exact combination of SNP genotypes comprising a signature and 0 if they did not. This term was included in the regression as an independent variable alongside covariates representing the top 5 genetic PCs (see Supplementary Table 5), with case-control status of the patients in the population (1 = case, 0 = control) as the dependent variable.

The limited number of patients with ICD-10 codes for long COVID provides very limited power to replicate, i.e. statistically validate individual disease signatures’ disease associations in AoU, especially given the need for false discovery rate correction when testing the many signatures identified in the Sano GOLD dataset. Instead, we began by testing whether we observed a statistically significant enrichment of disease signatures that are also positively correlated with long COVID in the AoU cohort after controlling for population substructure.

For each of the three sets of disease signatures identified in the original Sano GOLD study (Severe, Fatigue Dominant, and General), we first counted the number of signatures where the logistic regression returns a positive coefficient (i.e., odds ratio > 1) for the independent ‘genetic signature’ variable. Below we use the term ‘reproducing’ to denote signatures with odds ratio > 1 in the AoU test cohort, and ‘reproducibility rate’ to denote the percentage of tested signatures that have odds ratio > 1.

Some of the original signatures could not be evaluated in AoU because one or more of their component SNP genotypes are not included in the dataset. These were excluded from the analysis (see Supplemental Table 6). Most of these missing SNPs are represented on the Illumina GDA but we believe these data were likely filtered out during the AoU dataset’s QC processes.

Many of the long COVID disease signatures are non-independent due to shared component SNP genotypes and linkage disequilibrium, preventing us from using standard statistical tests to evaluate the significance of our observed reproducibility rates. We therefore used a permutation-based approach to generate the expected distribution of observed reproducibility rates under the null hypothesis (i.e., no association between signatures and disease). We randomly shuffled the case-control vector 100 times, reran the logistic regression analysis for every signature and counted the number of disease signatures that have odds ratio greater than 1 for each random permutation. The *p*-value of the observed results is equal to the number of permutations in which the number of signatures with odds ratios above 1 is greater than or equal to the number of signatures with odds ratios above 1 in the original analysis.

From previous experience with other diseases, we have found that reproducibility rates for disease signatures are positively correlated with the frequency of the signature in the population. To test whether this is true for long COVID we filtered each of the three sets of disease signatures from the original analysis to the subsets that occur in at least 4% or 5% of total cases in the AoU cohort (based on observations of reproducibility rates from prior unpublished studies in other diseases). We then reran the reproducibility analysis for each of these ‘high frequency’ subsets of signatures.

Finally, we evaluated the impact of signature complexity (i.e., number of SNP genotypes in the signature) on reproducibility rates. We split the set of signatures into sets comprised of 2, 3, 4, and 5 SNP genotypes and reran the analysis of reproducibility on each separately, with and without applying filtering by case-frequency.

### Ancestry-Specific Analyses

To test whether the observed rates of disease signature reproducibility broadly apply across traditionally under-served patient cohorts, we created three ancestry-specific sub-cohorts consistent with the demographic categories used to match cases with controls:

1. White – patients who self-identify as ‘white’
2. Black / African-American – patients who self-identify as ‘black’ and/or ‘African-American’
3. Hispanic / Latino - patients who self-identify as ‘Hispanic’, ‘Latino’, and/or ‘Latina’

Note that these cohorts are not all mutually exclusive, as AoU includes separate questionnaire questions for self-reported race and Hispanic/Latino identification.

We again used genetic principal components to control for any indirect relationships between signature frequency and disease prevalence resulting from population substructure (including relatedness or broader shared ancestry between patients). We conducted separate PCAs for each sub-cohort dataset using the approach described above for the whole cohort. We then selected the first five PCs as covariates for each ancestry-specific analysis after confirming that they explained sufficient variance in the dataset (see Supplementary Table 5).

We restricted the ancestry-specific analyses to the sets of signatures that occur in more than 5% of cases. Given the small sample size and low statistical power for ancestry-specific sub-cohorts, it is most appropriate to assess differences in reproducibility rate for the sets of signatures that exhibit the strongest reproducibility statistics in the full cohort.

### Evaluating Enrichment of Reproducing Long COVID Disease Signature in AoU Cohort

Finally, we tested whether any of the original disease signatures replicate, i.e., are individually significantly associated with long COVID in AoU. To minimize the FDR correction required for multiple tests, we restricted the analysis to the sets of high-frequency signatures that occur in more than 5% of cases. Output for the three original combinatorial analyses were assessed separately. Uncorrected *p*-values were obtained from the logistic regression with genetic PC covariates. ‘Reproducing’ signatures have *p*-values < 0.05 after FDR correction via the Benjamini-Hochberg procedure^43^. We also assessed significance using the more conservative Bonferroni adjustment^44^.

The SNPs in the disease signatures associated with long COVID in AoU were mapped to genes using an annotation cascade process against the human reference genome (GRCh38), as detailed in Das et al. (2022)^27^. SNPs located within the coding region of a gene (or genes) were mapped directly to the gene(s) and any remaining SNPs within 2kb upstream or 0.5kb downstream were mapped to the closest gene(s).

## Results

### Reproducibility of Overall Long COVID Disease Associations in AoU Cohort

We were able to test 5,343 of the 9,068 long COVID disease signatures originally identified in the three Sano GOLD sub-cohorts. The untested signatures all contained at least one SNP genotype that was not present in the post-QC AoU genotype dataset. Of the tested signatures, 1,766 occur in greater than 5% of cases and 3,100 occur in greater than 4% of cases in AoU.

When we restricted the analysis to signatures with case frequency greater than 5%, we consistently observed a significant enrichment of signatures (77% - 83%, *p* < 0.01) that are positively associated with long COVID in the AoU cohort, across all three sets of disease signatures (Table 3). Notably, the percentage of signatures with odds ratios greater than 1 in AoU is much larger than observed in any of the permutations where cases and controls were randomly assigned to patients (e.g., 82% vs. a maximum of 57% in the random permutations for the Severe cohort). This result confirms that many disease signatures are non-randomly associated with increased long COVID prevalence in AoU.

**Table 3.** Reproducibility statistics in AoU for long COVID disease signatures derived from three Sano GOLD sub-cohorts.

| Case Frequency Filter | Sub-Cohort | # signatures tested | % signatures with odds ratio > 1 in AoU | % signatures with odds ratio > 1 in permutations mean (range) | p-value |
| --- | --- | --- | --- | --- | --- |
| 5% | Severe | 109 | 82% | 42% (31% - 57%) | **<0.01** |
|  | Fatigue Dominant | 35 | 83% | 43% (23% - 71%) | **<0.01** |
|  | General | 1,622 | 77% | 44% (30% - 59%) | **<0.01** |
| 4% | Severe | 243 | 67% | 41% (30% - 56%) | **<0.01** |
|  | Fatigue Dominant | 85 | 71% | 43% (21% - 59%) | **<0.01** |
|  | General | 2,772 | 60% | 43% (30% - 59%) | **<0.01** |
| none | Severe | 668 | 35% | 43% (32% - 53%) | 0.96 |
|  | Fatigue Dominant | 740 | 34% | 46% (33% - 57%) | 0.98 |
|  | General | 3,935 | 44% | 42% (33% - 51%) | 0.36 |

Overall reproducibility rates are lower when we apply a less stringent 4% frequency cutoff, but the enrichment is still highly significant (60% - 71%, *p* < 0.01). That is, the observed number of signatures with odds ratios greater than 1 in AoU exceeds the maximum number of signatures with odds ratio greater than 1 in the random permutations. We did not observe any significant enrichment of reproducing signatures when we included low-frequency signatures in our analysis.

The distributions of odds ratios for the reproducing high-frequency (>5%) signatures are shown in Figure 1. 89% and 90% of the reproducing signatures from the Severe and Fatigue Dominant studies respectively have odds ratios greater than 1.1 in AoU, while 17% and 48% have odds ratios greater than 1.5. The mean odds ratio for the Severe signatures is 1.35 and the maximum is 2.09, while the mean odds ratio for the Fatigue Dominant signatures is 1.49 and the maximum is 2.22. Thus, the reproducing disease signal from these studies largely represents signatures that are individually strongly associated with increased disease prevalence.

**Figure 1.**
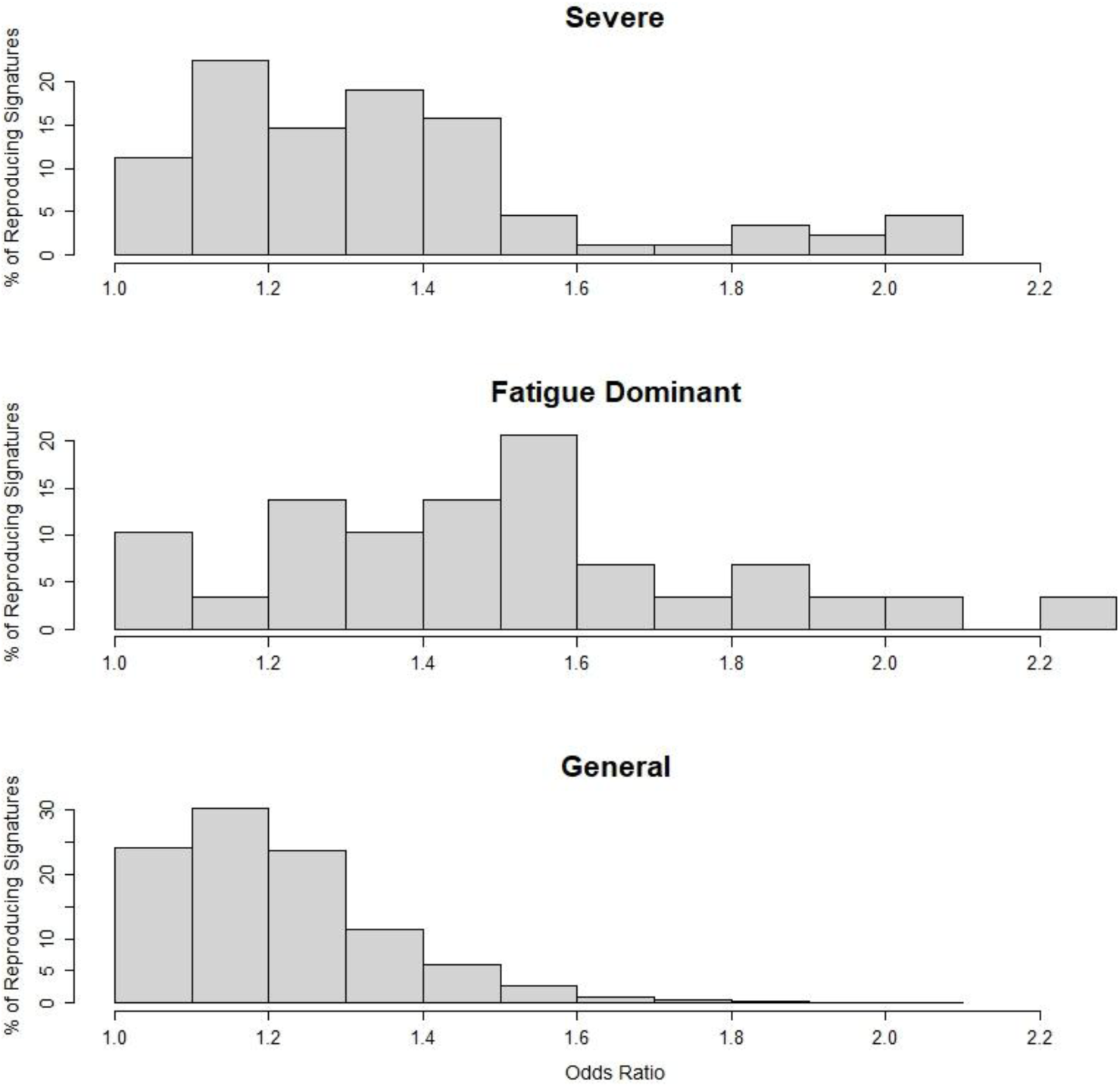
Distribution of observed odds ratios in AoU for reproducing signatures with high case frequencies (>5%).

The reproducing signatures from the General study tend to have lower odds ratios than the other studies (Figure 1). 75% of reproducing signatures have odds ratios greater than 1.1 in AoU and 5% have odds ratio greater than 1.5. The mean odds ratio is 1.21 and the maximum is 2.10. Thus, although these signatures included many with relatively weak disease associations, they also include signatures with strong effect sizes.

The relative enrichment of low odds ratios for signatures from the General study likely reflects the greater number of cases (Supplemental Table 4) and greater statistical power associated with the Sano GOLD study cohort. That is, the General study was better suited to detect signatures with lower effect sizes relative to the smaller Severe and Fatigue Dominant cohorts. The weaker disease associations may also reflect the relative reliability of the criteria used to define the Sano GOLD cohorts, as we believe that the case definition criteria for the General cohort is less accurate than the criteria used to identify patients with Severe and Fatigue Dominant long COVID subtypes.

Reproducibility statistics are strongest for high case frequency (>5%) signatures comprised of 4 or 5 SNP genotypes, as measured both by percent reproducing (i.e., odds ratio >1) and *p*-value (Table 4). Notably, across all three analyses, roughly twice as many 4- and 5-SNP signatures have odds ratios greater than 1 in AOU than would be expected due to random chance based on the mean reproducibility rates for the random permutations.

**Table 4.**
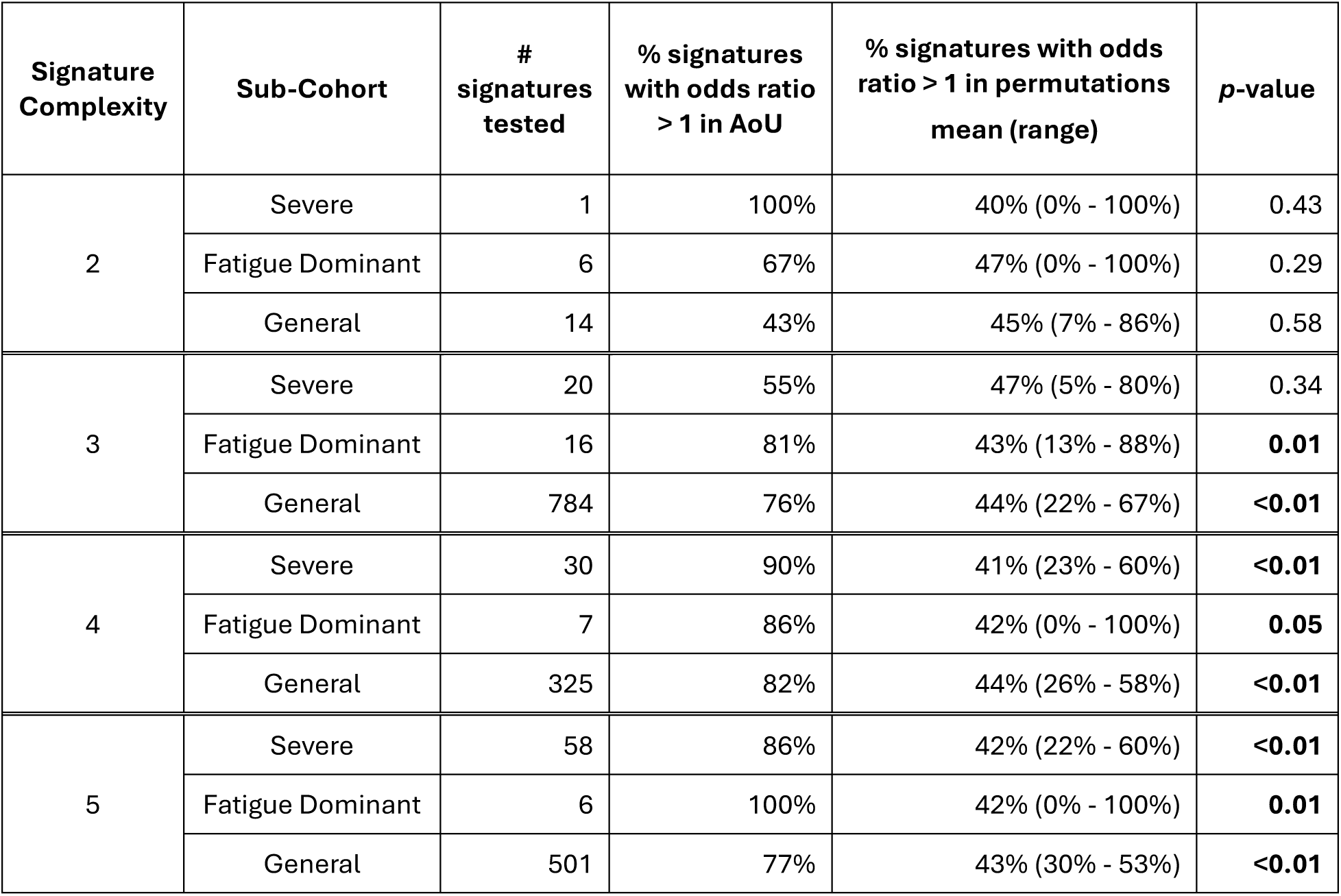
Reproducibility statistics by signature complexity (i.e., number of SNP genotypes comprising disease signatures) in AoU for high case frequency (>5%) long COVID disease signatures derived from three Sano GOLD sub-cohorts.

| Signature Complexity | Sub-Cohort | # signatures tested | % signatures with odds ratio > 1 in AoU | % signatures with odds ratio > 1 in permutations mean (range) | p-value |
| --- | --- | --- | --- | --- | --- |
| 2 | Severe | 1 | 100% | 40% (0% - 100%) | 0.43 |
|  | Fatigue Dominant | 6 | 67% | 47% (0% - 100%) | 0.29 |
|  | General | 14 | 43% | 45% (7% - 86%) | 0.58 |
| 3 | Severe | 20 | 55% | 47% (5% - 80%) | 0.34 |
|  | Fatigue Dominant | 16 | 81% | 43% (13% - 88%) | **0.01** |
|  | General | 784 | 76% | 44% (22% - 67%) | **<0.01** |
| 4 | Severe | 30 | 90% | 41% (23% - 60%) | **<0.01** |
|  | Fatigue Dominant | 7 | 86% | 42% (0% - 100%) | **0.05** |
|  | General | 325 | 82% | 44% (26% - 58%) | **<0.01** |
| 5 | Severe | 58 | 86% | 42% (22% - 60%) | **<0.01** |
|  | Fatigue Dominant | 6 | 100% | 42% (0% - 100%) | **0.01** |
|  | General | 501 | 77% | 43% (30% - 53%) | **<0.01** |

We similarly observed that reproducibility statistics are strongest for higher complexity signatures when applying a frequency cut-off of 4% (Supplemental Table 7). We observed no clear association between signature complexity and reproducibility rates for low frequency signatures (Supplemental Table 8).

To ensure that the sets of long COVID disease signatures are broadly reproducible across patients, we conducted separate analyses for self-reported white, black/African-American, and Hispanic/Latino sub-cohorts (Table 5).

**Table 5.**
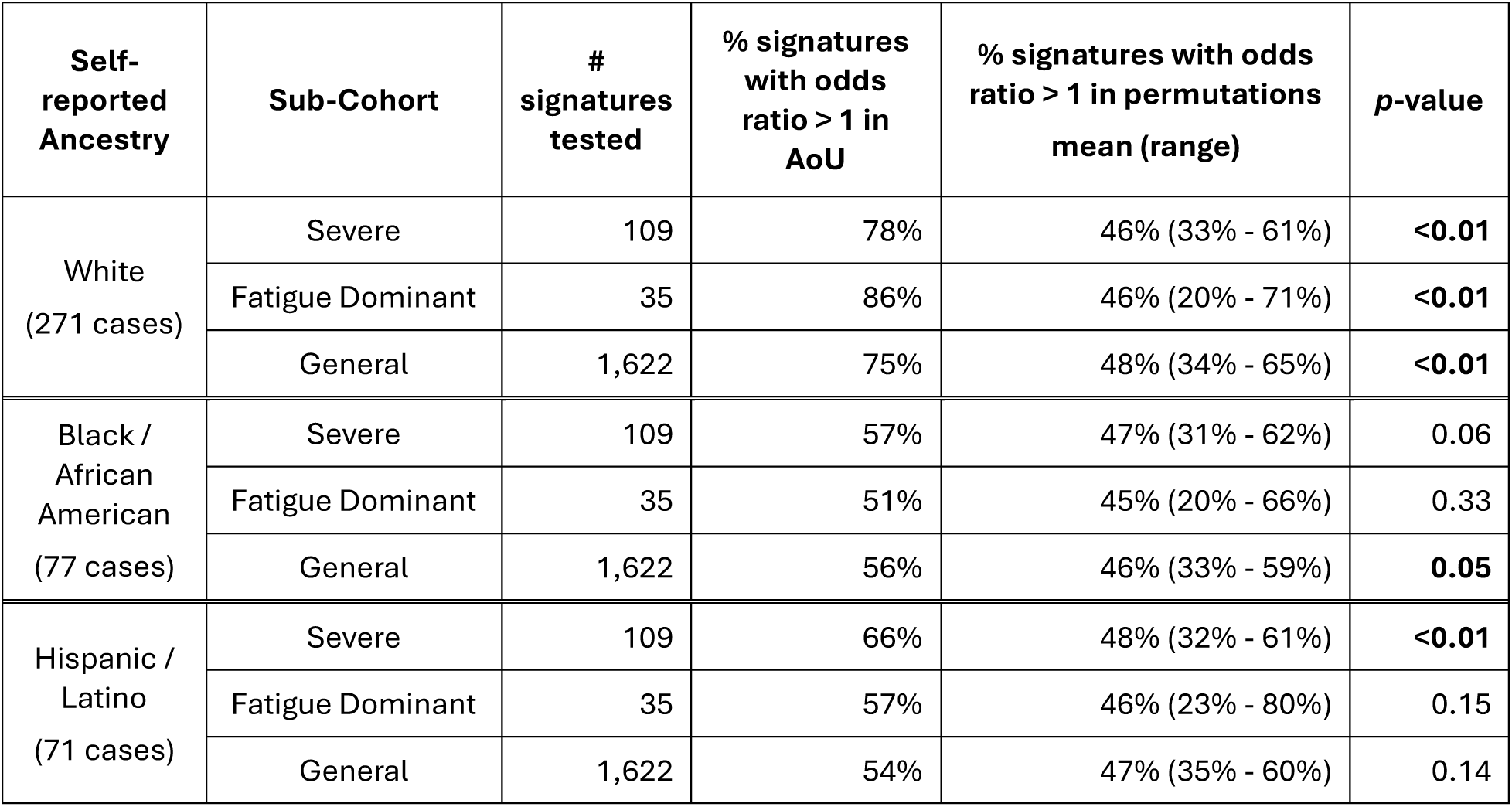
Reproducibility statistics in AoU for high case frequency long COVID disease signatures (>5% of cases) derived from three Sano GOLD sub-cohorts, broken down by self-reported ancestry.

We observed a highly significant enrichment of positively correlated disease signatures among self-reported white patients. This result confirms that the observed enrichment of reproducible disease associations in the all-ancestry cohort does not simply reflect population substructure in the dataset (i.e., indirect correlations between disease prevalence and signature frequency that arise due to shared correlations with ancestry).

Reproducibility rates were lower in the self-reported black/African-American and Hispanic/Latino sub-cohorts relative to the self-reported white sub-cohort, but consistently above the mean values observed in the random permutations. Two of the observed enrichment values were statistically significant (*p* < 0.05) despite the very small number of cases (71 and 77) and consequent weak statistical power in these sub-cohorts.

More than 85% of the long COVID genes identified across the three Sano GOLD long COVID cohorts mapped to one or more disease signatures that have >4% case frequency and were also positively associated with long COVID in the AoU cohort (see Table 6). Out of the 73 genes published in Taylor et al. (2023)^30^ 15 genes could not be tested due to missing SNPs in the AoU dataset. 76% (44/58) of the remining genes also map to disease signatures that reproduced in AoU. These genes are linked to a wide range of biological processes and mechanisms including dysregulated immune response and metabolic pathways, development of chronic inflammation, and cognitive dysfunction.

**Table 6.** Reproducibility statistics in AoU for genes associated with high case frequency (> 4% and >5%) long COVID disease signatures identified in three Sano GOLD sub-cohorts.

| Sub-Cohort | # Genes identified in Sano GOLD cohort | # Genes evaluated in AoU | # Genes mapped to signatures with >4% case frequency | # (%) of Genes mapped to signatures with >4% case frequency that have odds ratio >1 in AoU | # Genes mapped to signatures with >5% case frequency | # (%) of Genes mapped to signatures with >5% case frequency that have odds ratio >1 in AoU |
| --- | --- | --- | --- | --- | --- | --- |
| Severe | 43 | 35 | 35 | 30 (85.71%) | 25 | 22 (88%) |
| Fatigue Dominant | 35 | 28 | 16 | 15 (93.75%) | 6 | 5 (83.3%) |
| General | 165 | 151 | 150 | 140 (93.33%) | 138 | 128 (92.8%) |

Of the 13 repurposing gene candidates identified in Taylor et al. (2023), 11 (85%) map to at least one disease signature that reproduces in AoU (see Supplemental Table 9). These genes include *TLR4* which Taylor et al. (2023) noted has been shown to protect against long-term cognitive impairment pathology caused by SARS-CoV-2^45^. Inhibition of TLR4 in a mouse model was shown to prevent long term cognitive pathology including synapse elimination and memory deficits that are caused by the SARS-CoV-2 Spike protein. Previous clinical studies have shown that antagonizing TLR4 signaling has the effect of dampening the pathological cytokine storm observed in patients with severe acute COVID-19 and reduces mortality rates in hospitalized COVID-19 patients^46,47^.

### Replication of Individual Disease Signatures in All of Us

The above analyses focused on demonstrating an overall enrichment of disease signatures and genes that are positively correlated with long COVID in AoU, recognizing that the small size of the AoU cohort severely limits wide-scale replication. To achieve sufficient power to statistically validate individual signatures, we limited our replication analysis to the subset of signatures with case frequencies above 5%.

Four high-frequency disease signatures from the Severe Sano GOLD analysis were significantly associated with increased prevalence of long COVID in AoU, one of which was still significant after applying the more conservative Bonferroni FDR correction (Table 6). All four signatures are comprised of five SNP genotypes, each of which contributes to the overall association with disease in AoU (i.e., removing any of the SNP genotypes from the signature results in a lower odds ratio). This observation highlights the utility of the combinatorial analysis approach for identifying genetic disease associations.

Two of the replicating disease signatures from the Severe analysis mapped to the gene *CCDC146* and one mapped to *D2HGDH*. These genes have different functions and affect different potential mechanism of action hypotheses for their role in the development of long COVID. CCDC146 is a ubiquitous centriole and microtubule-associated protein linked to cognitive functioning and type 2 diabetes^48^. D2HGDH is an enzyme involved in mitochondrial functioning, also exhibits anti-inflammatory effects^49^.

Two disease signatures from the Fatigue Dominant Sano GOLD analysis were significantly associated with increased prevalence of long COVID in AoU, one of which was still significant after applying the more conservative Bonferroni FDR correction (Table 6). The latter is comprised of two SNP genotypes, while the other is comprised of five SNP genotypes. Each of the individual SNP genotypes contribute to the signatures’ association with disease in AoU.

None of the signatures from the General cohort in the Sano GOLD analysis were significantly associated with increased prevalence of long COVID in AoU. Although this output includes the signature most strongly associated with long COVID (by uncorrected *p*-value), it does not survive the stringent FDR correction for the large number of signatures from this analysis.

Finally, if we pool the signatures from the Severe and Fatigue Dominant cohorts into a single analysis (excluding the large number of signatures from the General cohort to avoid the need for stringent FDR correction), then 5 of signatures in Table 7 remain significant under the combined Benjamini-Hochberg FDR correction. These include all four significant signatures from the Severe analysis and the top signature from the Fatigue Dominant analysis.

**Table 7.** Replicating disease signatures that exhibit statistically significant associations with long COVID in AoU using Bonferroni-Hochberg FDR procedure. *p*-values in bold are also significant under Bonferroni FDR correction. Signatures from each cohort analysis were evaluated separately. Odds ratios reflect the number of total case and controls with genotype data for all component SNPs, which differs between signatures.

| Signature | # Cases | # Controls | Odds Ratio | *p*-value |
| --- | --- | --- | --- | --- |
| **Severe cohort** |  |  |  |  |
| (rs17035343 T/T)(rs1872513 C/C)(rs9312595 A/G)<br>(rs4936114 A/G)(rs12454570 C/C) | 30 | 149 | 2.09 | **0.0004** |
| (rs17035343 T/T)(rs1872513 C/C)(rs9312595 A/G)<br>(rs58438895 A/A)(rs4936114 A/G) | 29 | 144 | 2.09 | 0.0005 |
| (rs17035343 T/T)(rs1872513 C/C)(rs9312595 A/G)<br>(rs1109968 A/A)(rs4936114 A/G) | 29 | 144 | 2.09 | 0.0005 |
| (rs6716743 G/G)(rs2010874 T/T)(rs13096228 A/G)<br>(rs67844017 A/A)(rs79853277 T/T) | 28 | 136 | 2.15 | 0.001 |
| **Fatigue Dominant cohort** |  |  |  |  |
| (rs9515203 C/C)(rs11633336 A/A) | 27 | 120 | 2.33 | **0.0004** |
| (rs10914896 G/G)(rs10229643 A/A)(rs9960341 A/A)<br>(rs2076584 C/C)(rs17702926 C/C) | 23 | 117 | 2.02 | 0.003 |

## Discussion

Studies using traditional GWAS and meta-GWAS approaches on large patient populations (6,450 cases and 53,764 cases) respectively identified a single locus and three loci associated with long COVID^25,26^, although there was no statistical replication of the findings between these studies.

The original combinatorial analysis of Sano’s GOLD cohort identified 9,068 genetic disease signatures and 73 genes that were significantly enriched in two small UK-based long COVID patient cohorts (Severe n_cases=459 and Fatigue Dominant n_cases=477)^30^. In this original analysis, 28/43 genes found in the Severe cohort were also significantly associated with disease in the Fatigue Dominant cohort, and 25/35 genes from the original Fatigue Dominant analysis were also associated in the Severe cohort. 25 genes (15 from Severe and 10 from Fatigue Dominant) were found to be unique to those cohorts.

92% of the genes and 60-83% of the medium/high-frequency disease signatures from the Sano GOLD results that are also represented in the AoU dataset were positively correlated with long COVID in this independent US-based population. For disease signatures that occur in at least 5% of patients, between 77%-83% were positively correlated with long COVID prevalence in both the Sano GOLD and AoU cohorts, far more than we would expect to randomly observe if the signatures were uncorrelated with disease biology. Although we defined a ‘reproducing’ signature as one that has any odds ratio greater than 1, most reproducing signatures have relatively large odds ratios in AoU, indicating a strong association with increased disease prevalence.

At least five of the disease signatures found in Sano GOLD were individually significantly associated with increased prevalence of long COVID in the AoU population. The significant enrichment of positively-associated disease signatures further confirms that many additional signatures are non-randomly associated with disease but cannot be individually validated due to the very low statistical power provided by the small number of long COVID patients in the dataset (n=413). Together these results demonstrate a significant enrichment and reproduction of disease signal, broadly validating the results of the original analysis.

Importantly, the results of this paper provide strong supporting evidence for a much broader range of genetic associations with long COVID than has been uncovered by GWAS studies to date. This provides evidence highly consistent with a strong biological basis of the disease and the hypothesis that patients’ genetics influence their susceptibility to developing long COVID (and their predominant symptoms) following recovery from acute SARS-CoV-2 infection.

The AoU ancestry distribution differs significantly from the mainly (>91%) white British patient cohort used in the original combinatorial analysis. Disease signature reproducibility rates are very strong in the sub-cohort of self-identified white patients, as expected given the similarity in ancestry between that cohort with the original Sano GOLD dataset. Signature reproducibility rates are lower in sub-cohorts of self-identified black/African-Americans and Hispanic/Latinos, but we still observe significant enrichment of disease signatures despite very small sample sizes.

This therefore represents the first reproduction of long COVID genetic associations across multiple populations with substantially different ancestry distributions. Given the degree of reproducibility of results across diverse populations, these findings may have broad clinical application which could promote better health equity.

The lower signature reproducibility rates among the self-identified black/African American and Hispanic/Latino sub-cohorts relative to the self-identified white sub-cohort highlight a pressing need to identify large, diverse, well-phenotyped cohorts of long COVID patients. Many long COVID specific datasets such as Sano GOLD are comprised predominantly of patients with white European ancestry. In contrast, All of Us includes a highly diverse patient cohort, but lack of reliable data identifying which participants have a history of long COVID prevents us from reliably obtaining sufficient sample sizes to conduct a combinatorial analysis aimed at identifying novel disease signatures.

Having access to larger and more diverse populations with a confirmed diagnosis is essential to enabling primary analysis within these ancestry cohorts and adding to our understanding of the factors underpinning disease in those populations. In turn this would also help us build more inclusive and transferrable disease risk models.

Combinatorial analysis of diverse long COVID patient cohorts could potentially identify disease signatures that were not detected in predominantly white European cohorts due to low relative case frequencies, but which have greater frequency and importance for disease biology in other patient cohorts. Such signatures could be used to better estimate patients’ relative susceptibility to developing long COVID.

Alternatively, the disease signatures identified in the Sano GOLD cohort may have reduced effect sizes in non-white European cohorts due to an increased frequency of ‘actively protective’ signatures in those populations, i.e., one or more SNP genotypes that wholly or partially mitigate the disease associations of a set of ‘causative’ disease signatures^50^. The combinatorial analysis published in Taylor et al. (2023) focused only on causative disease signatures and did not include any analysis of protective signatures^30^ Incorporating actively protective features into the set of disease signatures should increase their predictivity for identifying ‘high-risk’ patients and improve reproducibility statistics.

### Evaluating the Output of the PrecisionLife Combinatorial Analysis Platform

We observed high rates of reproducibility among disease signatures derived from all the Sano GOLD cohorts and showed that these rates of disease signature reproducibility were strongly correlated with the frequency of signatures in the original study cohort. We observed slightly higher overall rates of reproducibility in the Severe and Fatigue Dominant cohorts which have fewer high case frequency disease signatures relative to the broader ‘General’ long COVID cohort.

Rates of reproducibility were highest for disease signatures comprised of four or five SNP genotypes, suggesting that combinatorial genetic interactions play an important role in the biology of long COVID. This also provides supporting evidence for the combinatorial analytic approach’s ability to detect a broad and clinically informative set of genetic disease associations in otherwise intractable complex diseases.

### The Predictive Value of Common vs Rare Signatures

In contrast to these mid/high case frequency signatures, when analyzing the entire set of disease signatures from the original analyses including low frequency signatures, only 34%-44% were consistently correlated with long COVID prevalence. This implies that rarer signatures may replicate between populations more poorly, an observation that is consistent with similar findings in GWAS and polygenic risk score studies^51,52,53,54,55^. There are a several explanations for this observed correlation between signature frequency and reproducibility rates.

First, statistical power is proportional to sample size, which is already limited in the reproducibility analysis due to the very small number of confirmed long COVID patients in AoU. Signatures with frequencies below 5% are expected to occur in 21 or fewer AoU cases. This small sample size results in large variance in expected rates of reproducibility under the null model and a high probability of observing odds ratios less than one due to random sampling even when signatures are biologically relevant to disease.

Second, due to the high case: control skew (1:10) in our dataset, rare signatures were often more likely to be negatively correlated with disease under the null model. In the most extreme scenario, a signature that occurs in one person in the dataset is 10 times more likely to randomly occur in a control (resulting in a negative odds ratio) than a case (resulting a positive odds ratio). This bias caused the mean numbers of signatures that randomly exhibit odds ratios above 1 in the null model permutations to range between 41%-46%, below the 50% expectation for a balanced dataset.

Third, rare signatures appeared to be more reflective of subpopulation structure in the original Sano GOLD dataset. Including genetic principal components as covariates resulted in 4% fewer high-frequency signatures (i.e., those that occur in >5% of total cases) that are positively correlated with long COVID, relative to a logistic regression that did not include covariates for population substructure. In contrast, including genetic principal components in the analysis resulted in 52% fewer replicating low-frequency signature (i.e., those that occur in <4% of total cases).

Finally, more complex disease signatures (i.e., those comprised of 4 or 5 SNP genotypes) generally occur at lower frequencies in the population simply because there are more possible genotype combinations for a larger set of SNPs. The risk of overfitting to a dataset is known to increase with tree depth when applying tree-based machine learning algorithms^56^ and the same potentially holds true for higher layer disease signatures derived from a layer-based mining approach. Applying a frequency filter therefore potentially mitigates the impacts of false positive SNPs by removing higher-order signatures.

We found no evidence, however, that increased signature complexity was associated with reduced reproducibility among high-frequency signatures. Rather, overall reproducibility rates were highest for 4-SNP and 5-SNP signatures relative to the small number of 2-SNP signatures. We also did not observe a correlation between signature complexity and reproducibility rate among low-frequency signatures. These results suggest that outputs of the combinatorial analyses of the Sano GOLD cohorts were not excessively overfitted to the original datasets and that presence of any false positive component SNP genotypes does not significantly affect the overall association with disease.

Although the results of this analysis suggests that false positive component SNP genotypes do not have a major effect on signature reproducibility, we could potentially improve the effect sizes and predictivity of these signatures by using AoU to further refine the set of signatures.

This step entails testing each signature individually and removing any component SNP genotype that does not enhance the signature’s association with disease in AoU. We have not included any refinement analysis in this study because it can potentially overfit the new set of signatures to the training dataset (AoU). A third cohort of long COVID patients would be required to properly evaluate the improvement in disease signature reliability that results from this refinement process.

### Limitations of Analysis

Reliably identifying which patients in AoU have a history of long COVID is currently challenging as we needed to rely on ICD-10 codes, which are known to be inconsistently and inaccurately applied, to identify known cases. As noted above, published estimates of long COVID prevalence in the United States range between 6.9% to 14%, yet fewer than 0.2% of individuals in AoU have ICD-10 codes associated with long COVID.

This suggests that many long COVID patients have not been assigned the appropriate ICD-10 code. As a result, more than 10% of the controls in our AoU study cohort potentially could represent misclassified cases with unreported long COVID.

This type of phenotypic misclassification in datasets will generally weaken the observed effect sizes by artificially inflating the similarity between cases and controls^57^. This behavior is potentially problematic for reproducibility analyses, as the dilution of signal decreases the statistical power of the analysis^58^. For example, phenotypic misclassification increases the probability that a signature that is biologically correlated with increased disease risk will nonetheless exhibit an odds ratio less than 1 due to random sampling effects.

We therefore expect that the high degree of phenotypic misclassification in our dataset will have worked to reduce the overall rates of observed signature reproducibility. As such, the reproducibility statistics presented in this paper probably represent a low-end estimate of the true reproducibility rate.

### Applications for healthcare

The identification of a set of genetic signatures that are consistently associated with increased prevalence of long COVID offers many opportunities for improving treatment of this poorly understood but highly prevalent and debilitating disease.

Firstly, although we have insufficient power to validate the full set of individual signatures in AoU, demonstrating that reproducing signatures are significantly enriched in a second dataset provides important confirmatory evidence of the original findings of the combinatorial analytics approach. To provide insights into potential drug therapies for long COVID, we further tested whether the signatures associated with novel drug targets and their related drug repurposing candidates are significantly correlated with increased long COVID prevalence in AoU. 27/30 (90%) of the genes represented in the >5% disease signatures and 11 out of the 13 drug repurposing candidates identified in the original study were reproduced in this study. This lends weight to their prioritization in clinical efficacy trials especially for those with generic drugs.

Controlled open-label studies of similar design to the RECOVERY trial in Covid-19, which rapidly identified dexamethasone as an effective frontline therapy^59^, can be undertaken on this set of generic drugs, benefiting from the additional evidence that one or more selected therapies is more likely to help the subset of patients who have those mechanisms’ disease signatures in their genetic makeup.

Secondly, we can use the insights into disease biology that are reflected by the reproducing disease signatures to construct a combinatorial risk score that evaluates an individual patient’s relative genetic susceptibility towards developing long COVID. Although genetic risk scores are not strictly diagnostic, especially in a pathogen triggered disease, they have substantial potential to be used by physicians for differential triage, i.e., to rapidly gauge the relative likelihood of different diagnoses when presented with ambiguous or indistinct symptoms and refer patients and/or select treatment options accordingly. As the utilization of large-scale COVID-19 testing fades, alternative tests that can help differentiate patients with long COVID from patients with other illnesses with similar symptoms will become increasingly useful in healthcare settings.

Constructing a combinatorial risk score from disease signatures is a more complex challenge than a conventional polygenic risk model – the latter assumes that all features (SNPs) act independently, whereas combinatorial disease signatures are often inherently correlated due to the sharing of SNP genotypes. Machine learning approaches can disentangle this complexity and non-independence and combine features such as disease signatures and their component SNP genotypes into a single predictive model. Although the small sample size of the AoU dataset is sufficient to train a combinatorial risk score using machine learning, a third (currently unavailable) independent dataset would be required to properly evaluate the relative increase in long COVID prevalence between subsets of patients flagged as having high and low genetic susceptibility.

Finally, the set of replicating disease signatures can be used to mechanistically stratify patients based on the causative etiologies most likely associated with their form of long COVID. This first entails assigning disease signatures to one or more mechanism-of-action (MoA) cluster/s based on the gene(s) associated with the component SNP genotypes. We can then assess whether a patient has a significant excess or lack of disease signatures associated with a given MoA relative to the distribution of signature counts in the larger long COVID community. In essence, this mechanistic stratification tool is comprised of multiple combinatorial risk scores, each for a different set of mechanism related disease signatures. This can provide insight in the clinic into the selection of therapies that are matched to a patient’s personal genetic makeup.

Unlike standard risk scores, which can be used to inform public health applications but provides more limited utility for personalized precision medicine^60^, a mechanistic stratification tool would potentially ultimately enable healthcare practitioners to identify individualized treatment regimens including single- or multi-drug therapies that are most likely to generate a positive outcome for a given patient. In the case of long COVID these mechanistic insights also have other potential applications, as the Taylor et al. (2023) combinatorial analysis also found evidence for substantial overlap in disease biology between long COVID and myalgic encephalitis / chronic fatigue syndrome (ME/CFS)^30^.

### Conclusion

The level of reproducibility of results from the original Sano GOLD long COVID study in the All of Us population to the extent demonstrated is highly encouraging for the study of long COVID and other similarly complex diseases. These findings redefine our understanding of long COVID by uncovering a broad spectrum of reproducible genetic signatures, laying the foundation for new diagnostic innovations and targeted therapies that have the potential to revolutionize care for millions suffering from this debilitating condition worldwide.

The study demonstrates the level of reproducibility of results achievable using combinatorial analysis, even across very small populations with diverse ancestries in highly heterogenous diseases. Increasing reproducibility across patients with different ancestries is critically important for improving equitable representation and access to healthcare solutions. All of these studies would nonetheless obviously benefit from larger datasets with a wider population diversity, more secure diagnosis, more harmonized health/symptom surveys and deeper genomic, longitudinal clinical, immunological and metabolic data.

The results provide further compelling evidence for the detailed description of the genetic components of long COVID’s complex disease biology that was presented in the original combinatorial analysis study^30^. We hope that this will provide confidence to explore some of these mechanisms and drug targets and help advance research into novel ways to diagnose the disease and accelerate the discovery and selection of better therapeutic options, both in the form of newly discovered drugs and/or the immediate prioritization of coordinated investigations into the efficacy of repurposed drug candidates.

We also hope that these findings will better establish a stronger appreciation of the role of genetic contributions to the etiology and lived experience of disease in long COVID patients and prove its underlying biological basis to the clinical community.

For the first time, a definitive test for the disease would enable clinicians to rapidly and accurately identify and triage patients, ensuring they receive timely and equitable access to care, and reducing the potential for misdiagnosis. It would also establish a definitive framework for measuring the public health impact of the disease, informing health policy and helping strategically prioritize research initiatives to make more rapid progress in addressing this massive global challenge and improve patients’ lives.

## Data Availability

Only data from existing All of Us and Sano GOLD study cohorts were analyzed and no new source data were collected for this study. Aggregate-level data for the All of Us cohort is publicly available at https://databrowser.researchallofus.org/ (Public Tier dataset). Individual-level data for the All of Us cohort, available in the Controlled Tier dataset, can be analyzed by approved researchers on the Researcher Workbench.

## Acknowledgements

Research described in this article has been conducted using data from the All Of Us Research Program and Sano Genetics’ Long COVID GOLD study. We gratefully acknowledge All of Us and Sano Genetics’ Long COVID GOLD participants for their contributions, without whom this research would not have been possible. We also thank the National Institutes of Health’s All of Us Research Program and Sano Genetics for making available the participant data examined in this study. Special thanks to Gert Møller and Claus Erik Jensen, who initially developed the combinatorial analytics methodology, and the rest of the PrecisionLife and Metrodora teams.

## Author contributions

SG, AR, MS, JS, SD, RG and KT contributed to the design of the study. JS designed the reproducibility analyses. MP, KC, JS, and SD performed the analyses described in this manuscript. MP and KC conducted the analyses on the Researcher Workbench. All authors contributed to writing the manuscript and consent to publication.

## Funding

The project was funded entirely by Metrodora Foundation and PrecisionLife Ltd.

## Ethics approval and consent to participate

The Sano GOLD study has approval from the Wales Research Ethics Committee (REC) (IRAS 291221). Consent to participate has been received from all participants. Institutional Reviewing Board (IRB) approval was obtained prior to enrollment of patients in the All of Us Research Program. Informed consent for all participants is conducted in person or through an eConsent platform that includes primary consent, HIPAA Authorization for Research use of EHRs and other external health data, and Consent for Return of Genomic Results. The protocol was reviewed by the Institutional Review Board (IRB) of the All of Us Research Program (IRB Approval Date: Dec 03, 2021). The All of Us IRB follows the regulations and guidance of the NIH Office for Human Research Protections for all studies, ensuring that the rights and welfare of research participants are overseen and protected uniformly. The All of Us Research Program is supported by the National Institutes of Health, Office of the Director: Regional Medical Centers (OT2 OD026549; OT2 OD026554; OT2 OD026557; OT2 OD026556; OT2 OD026550; OT2 OD 026552; OT2 OD026553; OT2 OD026548; OT2 OD026551; OT2 OD026555); Inter agency agreement AOD 16037; Federally Qualified Health Centers HHSN 263201600085U; Data and Research Center: U2C OD023196; Genome Centers (OT2 OD002748; OT2 OD002750; OT2 OD002751); Biobank: U24 OD023121; The Participant Center: U24 OD023176; Participant Technology Systems Center: U24 OD023163; Communications and Engagement: OT2 OD023205; OT2 OD023206; and Community Partners (OT2 OD025277; OT2 OD025315; OT2 OD025337; OT2 OD025276). Results reported are in compliance with the All of Us Data and Statistics Dissemination Policy disallowing disclosure of group counts under 20 to protect participant privacy.

## Declaration of Competing Interest

AR is an employee of Metrodora Foundation, SG and RG are co-chairs of Metrodora Foundation’s Scientific Advisory Board. JS, SD, KT, KC, MP and SG are employees of PrecisionLife Ltd. S.G. is a shareholder of PrecisionLife, Ltd.

