## Supplementary Data for "Reproducibility of Genetic Risk Factors Identified for Long COVID using Combinatorial Analysis Across US and UK Patient Cohorts with Diverse Ancestries"

**Supplemental Table 1.** Exclusionary criteria for long COVID controls. Any patient that was assigned to one or more of the following All of Us (AoU) "concepts" (accessed on 10$^{th}$ December 2024) was excluded from the list of potential controls. Participant counts below 20 are displayed as 20.

| **Searchable AoU Concept** | **AoU participant count** |
|---|---:|
| Post COVID-19 condition (ICD-10 U09.9) | 520 |
| Fatigue | 60,860 |
| Chronic fatigue syndrome | 8,600 |
| Malaise or fatigue | 7,300 |
| Postviral fatigue syndrome | 380 |
| Postural orthostatic tachycardia syndrome (POTS) | 80 |
| Muscle fatigue | 40 |
| Accommodative fatigue | 20 |
| Tinnitus | 17,660 |
| Subjective tinnitus | 2,660 |
| Noises in ear | 20 |
| Tinnitus of vascular origin | 700 |
| Leudet's tinnitus | 20 |
| Tinnitus arising from tensor tympani | 20 |
| Tinnitus of muscular origin | 20 |

**Supplemental Table 2.** Demographic breakdown of cases and controls in the final All of Us long COVID cohort.

| Category | Value | Cases (n=413) | Controls (n=4130) |
|---|---|---|---|
| | Age [median (IQR)] | 60 (47-70) | 58 (43-69) |
| Sex | F | 69.7% | 68.3% |
| Sex | M | 30.3% | 31.7% |
| Self-reported race | Asian or None Indicated | 15.7% | 17.0% |
| Self-reported race | Black or African American | 18.6% | 19.5% |
| Self-reported race | White | 65.6% | 63.5% |
| Self-reported Ethnicity | Hispanic or Latino | 17.2% | 18.5% |
| Self-reported Ethnicity | Not Hispanic or Latino | 82.8% | 81.5% |
| Co-morbidities | Asthma (J45) | 39.0% | 19.0% |
| Co-morbidities | Chronic ischemic heart disease (I25) | 20.1% | 11.9% |
| Co-morbidities | Diabetes type 2 (E11) | 31.5% | 22.8% |
| Co-morbidities | Irritable bowel syndrome (K58) | 12.8% | 4.5% |
| Co-morbidities | Chronic kidney disease (N18) | 17.0% | 10.9% |
| Co-morbidities | Rheumatoid arthritis (M05 + M06) | 7.5% | 3.9% |
| Co-morbidities | Chronic fatigue syndrome (concept) | 14.3% | - |

**Supplemental Table 3.** Comparison of self-reported demographic data with predicted genetic ancestry distribution[1] of individuals within the All of Us (AoU) long COVID cohort. The genetic ancestry predictions were only available for participants who had whole genome sequencing (WGS) data in AoU.

| **Self reported demographics** | **Demographic category** | **Total number of individuals with predicted genetic ancestry in AoU** | **Matching Predicted Genetic ancestry[1]** | **Individuals with matching predicted genetic ancestry** | **% of individuals with matching genetic ancestry** |
|---|---|---|---|---|---|
| Race | Asian/None Indicated | | | | |
| Race | Black or African American | 705 | African ancestry (AFR) | 701 | 99.4% |
| Race | White | 2334 | European ancestry (EUR) | 2263 | 97.0% |
| Ethnicity | Hispanic or Latino | 700 | Latino/admixed American ancestry (AMR) | 617 | 88.1% |
| Ethnicity | Not Hispanic or Latino | | | | |

**Supplemental Table 4.** Summary of the three long COVID cohorts that were derived from GOLD study.

| **Long COVID cohort derived from GOLD study** | **Case Selection Criteria** | **Control Selection Criteria** | **Case count** | **Control count** | **Count of Disease signatures identified by combinatorial analysis** |
|---|---|---|---|---|---|
| **Severe** | Total Change score was calculated using the difference in self-reported scores pre- and post-acute COVID-19 for three symptom groups –respiratory, fatigue and mental health. Individuals with a 'Total Change' score greater than the upper quartile of the distribution | Individuals with a 'Total Change' score greater than or equal to 0 but below the median of the distribution | 459 | 864 | 1,188 |
| **Fatigue Dominant** | Fatigue Change score was calculated using the difference in self-reported scores relating to fatigue symptoms pre- and post-acute COVID-19. Individuals with a 'Fatigue Change' score greater than the upper quartile of the distribution. | Individuals with a 'Fatigue Change' score greater than or equal to 0 but below the median of the distribution. | 477 | 909 | 1,435 |
| **General** | Individuals with recovery time more than 12 weeks since COVID-19 diagnosis | Individuals who recovered within 12 weeks since COVID-19 diagnosis | 703 | 698 | 6,455 |

**Supplemental Table 5.** Principal component analysis (PCA) eigenvalue for All of Us long COVID dataset and self-reported ancestry sub-cohorts. Top 5 PCs were selected to represent population substructure in all datasets.

| Principal Component | Full Cohort Eigenvalues | Self-reported White Eigenvalues | Self-reported Black / African-American Eigenvalues | Self-reported Hispanic / Latino Eigenvalues |
|---|---|---|---|---|
| **PC1** | **291.96** | **8.15** | **5.72** | **24.96** |
| **PC2** | **50.52** | **4.00** | **1.55** | **9.10** |
| **PC3** | **7.40** | **2.35** | **1.52** | **1.55** |
| **PC4** | **2.42** | **2.03** | **1.39** | **1.49** |
| **PC5** | **2.14** | **1.60** | **1.38** | **1.44** |
| PC6 | 1.82 | 1.55 | 1.37 | 1.40 |
| PC7 | 1.78 | 1.48 | 1.36 | 1.40 |
| PC8 | 1.72 | 1.47 | 1.35 | 1.37 |
| PC9 | 1.72 | 1.44 | 1.35 | 1.36 |
| PC10 | 1.70 | 1.42 | 1.34 | 1.36 |
| PC11 | 1.69 | 1.41 | 1.33 | 1.34 |
| PC12 | 1.69 | 1.41 | 1.32 | 1.34 |
| PC13 | 1.68 | 1.41 | 1.31 | 1.34 |
| PC14 | 1.67 | 1.41 | 1.31 | 1.33 |
| PC15 | 1.66 | 1.41 | 1.31 | 1.32 |
| PC16 | 1.65 | 1.41 | 1.30 | 1.32 |
| PC17 | 1.64 | 1.40 | 1.30 | 1.31 |
| PC18 | 1.63 | 1.40 | 1.29 | 1.31 |
| PC19 | 1.63 | 1.40 | 1.29 | 1.30 |
| PC20 | 1.62 | 1.40 | 1.28 | 1.30 |

**Supplemental Table 6.** Summary of long COVID SNPs and disease signatures from the Sano GOLD study that could be mapped to All of Us (AoU) long COVID genotype array data.

| Long COVID cohort derived from GOLD study | Total number of disease signatures | SNP count in disease signatures | Count of SNPs missing in AoU genotype dataset | % SNPs mapped in AoU genotype dataset | No. of disease signatures with missing SNPs | No. of disease signatures mapped in AoU | % of disease signatures mapped in AoU |
|---|---|---|---|---|---|---|---|
| Severe | 1188 | 2321 | 158 | 93.2% | 520 | 668 | 56.2% |
| Fatigue Dominant | 1435 | 2679 | 224 | 91.6% | 695 | 740 | 51.6% |
| General | 6445 | 7556 | 663 | 91.2% | 2510 | 3935 | 61.1% |

**Supplemental Table 7.** Reproducibility statistics by signature complexity (i.e., number of SNP genotypes comprising signatures) in All of Us for moderate-high case-frequency (>4%) long COVID disease signatures derived from three Sano Genetics Gold sub-cohorts.

| # SNP-genotypes in signature | Cohort | # signatures tested | % signatures odds ratio > 1 in AoU | % signatures odds ratio > 1 in permutations Mean (range) | *p*-value |
|---|---|---|---|---|---|
| 2 | Severe | 1 | 100% | 35% (0% - 100%) | 0.35 |
| | Fatigue Dominant | 12 | 58% | 47% (8% - 92%) | 0.35 |
| | General | 15 | 40% | 45% (7% - 87%) | 0.69 |
| 3 | Severe | 37 | 32% | 43% (5% - 80%) | 0.89 |
| | Fatigue Dominant | 40 | 65% | 42% (13% - 88%) | 0.02 |
| | General | 1040 | 62% | 45% (22% - 67%) | 0.06 |
| 4 | Severe | 70 | 73% | 41% (24% - 54%) | <0.01 |
| | Fatigue Dominant | 14 | 86% | 42% (0% - 86%) | 0.02 |
| | General | 597 | 63% | 43% (30% - 63%) | 0.01 |
| 5 | Severe | 136 | 73% | 41% (23% - 60%) | <0.01 |
| | Fatigue Dominant | 19 | 79% | 41% (11% - 79%) | 0.01 |
| | General | 1122 | 57% | 42% (31% - 56%) | <0.01 |

**Supplemental Table 8.** Reproducibility statistics by signature complexity (i.e., number of SNP genotypes comprising signatures) in All of Us for all long COVID disease signatures derived from three Sano Genetics Gold sub-cohorts.

| # SNP-genotypes in signature | Cohort | # signatures tested | % signatures odds ratio > 1 in AoU | % signatures odds ratio > 1 in permutations Mean (range) | *p*-value |
|---|---|---|---|---|---|
| 2 | Severe | 2 | 50% | 43% (0% - 100%) | 0.66 |
| | Fatigue Dominant | 21 | 33% | 44% (14% - 81%) | 0.84 |
| | General | 15 | 40% | 45% (13% - 93%) | 0.66 |
| 3 | Severe | 58 | 24% | 43% (29% - 62%) | 1.00 |
| | Fatigue Dominant | 152 | 35% | 44% (31% - 58%) | 0.87 |
| | General | 1118 | 58% | 42% (26% - 67%) | 0.12 |
| 4 | Severe | 163 | 37% | 41% (30% - 55%) | 0.77 |
| | Fatigue Dominant | 117 | 30% | 47% (29% - 67%) | 0.99 |
| | General | 878 | 44% | 42% (30% - 66%) | 0.35 |
| 5 | Severe | 136 | 73% | 36% (30% - 58%) | 1.00 |
| | Fatigue Dominant | 19 | 79% | 34% (31% - 58%) | 0.95 |
| | General | 1122 | 57% | 36% (33% - 54%) | 0.93 |

**Supplemental Table 9.** Genes identified as repurposing candidates in the Sano long COVID study published in Taylor et al (2023)[2] that were reproduced in the All of Us (AoU) long COVID study.

| **Gene** | **Protein class** | **PHAROS target classification** | **Reproducibility in AoU Sub-Cohort (case frequency of signatures)** |
|---|---|---|---|
| CETP | Transporter | Tchem | Severe (> 4%) |
| GUCY1A2 | Enzyme | Tclin | Severe (> 4%) |
| HPN | Enzyme | Tchem | Fatigue Dominant (> 4%) |
| MAPK9 | Enzyme | Tchem | Fatigue Dominant (> 4%) |
| PDE4D | Enzyme | Tclin | Severe (> 4%) |
| POR | Enzyme | Tbio | Fatigue Dominant (> 4%) |
| PRODH | Enzyme | Tbio | Severe (> 4%) |
| RRBP1 | | Tbio | Fatigue Dominant (> 4%) |
| SLC12A1 | Transporter | Tclin | Fatigue Dominant (> 4%) |
| TLR4 | Enzyme, Transporter | Tchem | Severe (> 4%) |
| TNIK | Enzyme | Tchem | Fatigue Dominant (> 4%) |
